# A time series forecasting of the proportion of SARS-CoV-2 N501Y lineage in North America

**DOI:** 10.1101/2021.03.30.21254648

**Authors:** Elena Quinonez, Majid Vahed, Abdolrazagh Hashemi Shahraki, Mehdi Mirsaeidi

**Author notes:** Corresponding Author: Mehdi Mirsaeidi, 1951 NW 7th Ave # 2235, Miami, FL 33136.

## Abstract

**Introduction:** The outbreak of pneumonia known as SARS-COV-2 and newly-emerging South African (B.1.351), the United Kingdom (B.1.1.7) and Brazil (P.1) variants have led to a more infectious virus and potentially more substantial loss of neutralizing activity by natural infection or vaccine-elicited antibodies.

**Methods:** We identified prevalent mutations using the spike receptor-binding domain (S-RBD) of SARS-CoV-2 deposited in the Nextstrain global database and comparing them to the Wuhan-Hu-1/2019 genomic sequence as a reference. Then we calculated the percentages of mutant genomes from the total regional subsample isolates from December 2019 to the end of January 2021. We developed two separate time series forecasting models for the SARS-CoV-2 B.1.1.7 variant. The computational model used the structure of the S-RBD to examine its interactions with the neutralizing antibody, named CV30 (isolated from a patient), and human angiotensin-converting enzyme 2 (hACE-2), based on a hybrid algorithm of template-based modeling to predict the affinity of S protein to the neutralizing antibodies and hACE-2 receptor.

**Results:** The proportion of the B.1.1.7 strain in North America is growing fast. From these computations, it seems that the S-RBD and hACE-2 proteins are less favorable for the South African strain (K417N, E484K, and N501Y) as compared to the wild type structure and more favorable for B.1.1.7 and P.1 variants. In the present of crystallized CV30 neutralizing antibodies, docking scores suggest antibodies can be partially neutralize the B.1.1.7 variant, and, less efficiently, the B.1.351 and P.1 variants.

**Conclusion:** The rapid evolution of SARS-CoV-2 has the potential to allow the newly-emerged B.1.351, and P.1 variants to escape from natural or vaccine-induced neutralizing immunity and viral spreading.

## Background

The family Coronaviridae contains important zoonotic pathogens such as SARS-CoV1, MERS-COV and SARS-CoV2, that can cause mild to severe respiratory infections in humans ^1^. These virions are among the largest 1 RNA viruses and are characterized by their roughly spherical shape with large spike receptor-binding domain S-RBD glycoproteins that extend 16-21 nm from the viral envelope. The S-RBD of SARS-CoV2, which interacts with the human angiotensin-converting enzyme 2 (hACE-2) to mediate entry into cells ^2,3^, has been the main target for vaccine development ^4^. Routine surveillance of the genomic profile of the SARS-CoV-2 is crucial to discovering relationships between developing viral mutations and transmission rates, vaccine efficacy, and epidemiological tracing.

Several newly-emerging variants are circulating the world, raising concerns about their impact on infectivity and mortality, as well as the effectiveness of current vaccines. In South Africa (S.A), a new variant of SARS-CoV-2 (known as 501Y.V2 or B.1.351) has emerged which carries three mutations at important locations in the S-RBD (K417N, E484K and N501Y) ^5^. The B.1.351 variant represents approximately 36% of the African subsample ^6^. Evolution of B.1.351 resulted to more than16-fold increase of COVID-19 incidence in Zambia ^7^. The possibility of a similar experience in the U.S and other countries is a real threat ^8^. A second new variant, known as 501Y.V3 or P.1, carries three mutations in the S-RBD domain (K417T, E484K, and N501Y) was detected for the first time in Brazil ^9^ and then in other countries including the U.S ^10,11^. A third variant, called 202012/01 (also known as 501Y.V1 or B.1.1.7) has been identified in the United Kingdom (U.K.) ^12^, and soon after became the primary emerging variant in many countries^13-18^. Epidemiological analysis has shown that B.1.1.7 is highly transmissible (50 to 70% more) than other SARS-COV-2 variants ^12,19^. The N501Y mutation is shared between all three variants (Brazil: P.1, S.A: B.1.351, and U.K: B.1.1.7) but the later lineage (B.1.1.7 variant) has several additional ‘signature’ mutations in the SARS-CoV-2 S protein, including deletions at 69-70 and 144, as well as mutations A570D, D614G, P681H, T716I, S982A, and D1118H ^20-22^. The emergence of mutations in position 501 is of major concern because it involves one of the six key amino acid residues determining a tight interaction of the S-RBD with its receptor (hACE-2) ^22^. In the U.S., the Centers for Disease Control (CDC) recently predicted that the number of patients with the B.1.1.7 variant will be increasing ^18^ but did not show the trend of the proportional increase of this mutation and the potential underlying mechanism. We report here the forecast of B.1.1.7 variant isolation in the next 12 months with two vaccination efficacy scenarios in the U.S. We also predict the binding efficacy of neutralizing CV30 antibody and hACE-2 receptor in these new mutant variants.

## Results

SARS-CoV2 strain B.1.351 was tracked for the first time in South Africa in late August 2020, a few days before the emergence of P.1 in Brazil (Fig.1). Our analysis also shows that B.1.1.7 emerged in the U.K. in early December 2020 (Fig.1). More interestingly, the first mutation at position of 501 and 484 had been isolated from Côte d’Ivoire; West Africa. Tracking the variants in the U.S also revealed that B.1.1.7 has emerged at the end of 2020 (late December), while B.1.351 and P.1 both emerged at almost same time, the end of January 2021 (Fig.1).

**Figure 1.**
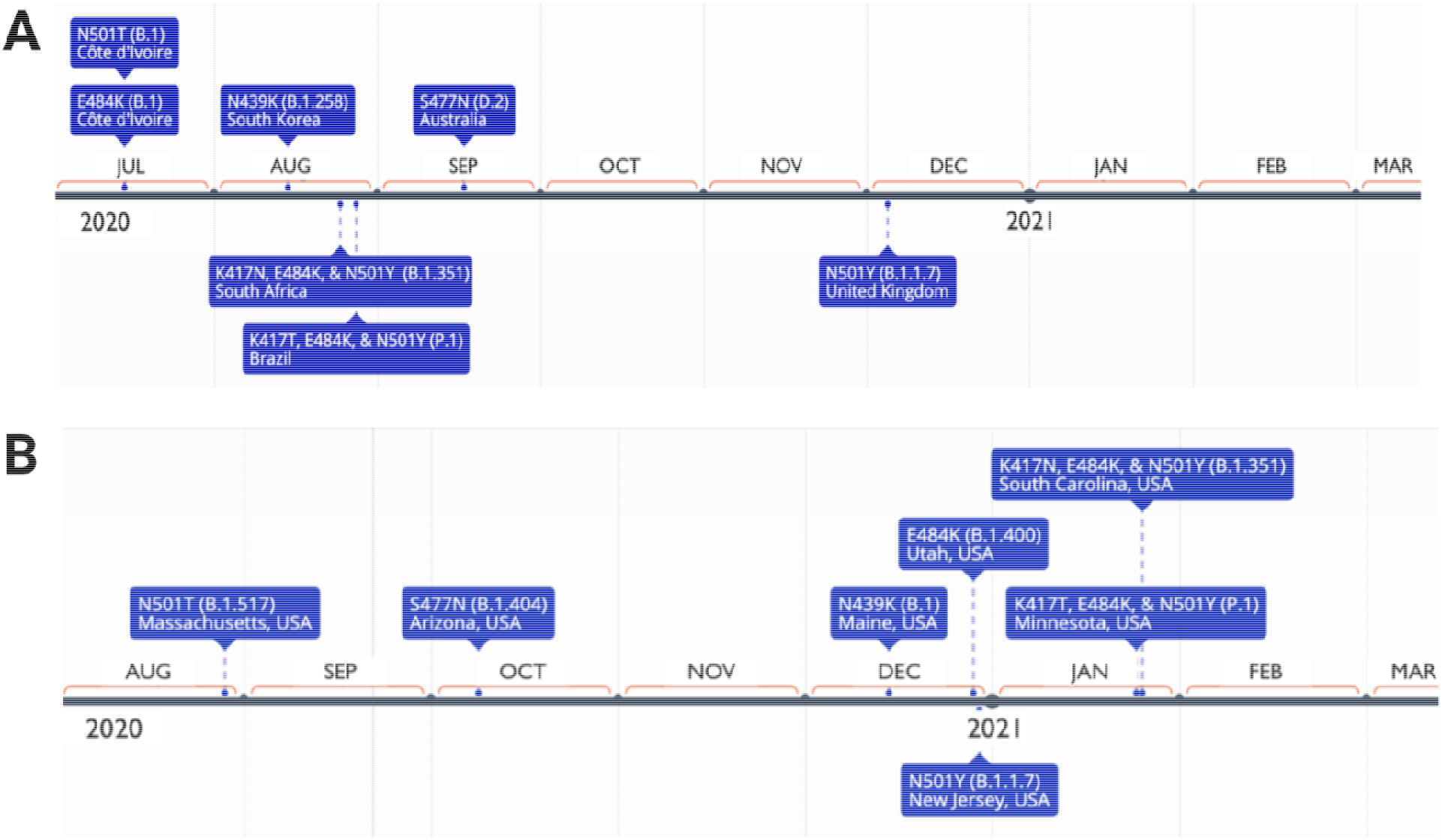
The timeline of RBD mutations in the world (above), and in the U.S. (below).

Forecast analysis data for models 1 and 2 are shown in Fig.2. Per model 1, until Oct 2021, all isolates from the patients with COVID19 will carry B.1.1.7 strain mutations. However, in model 2, almost 25% of isolated samples will exhibit this mutation.

**Figure 2.**
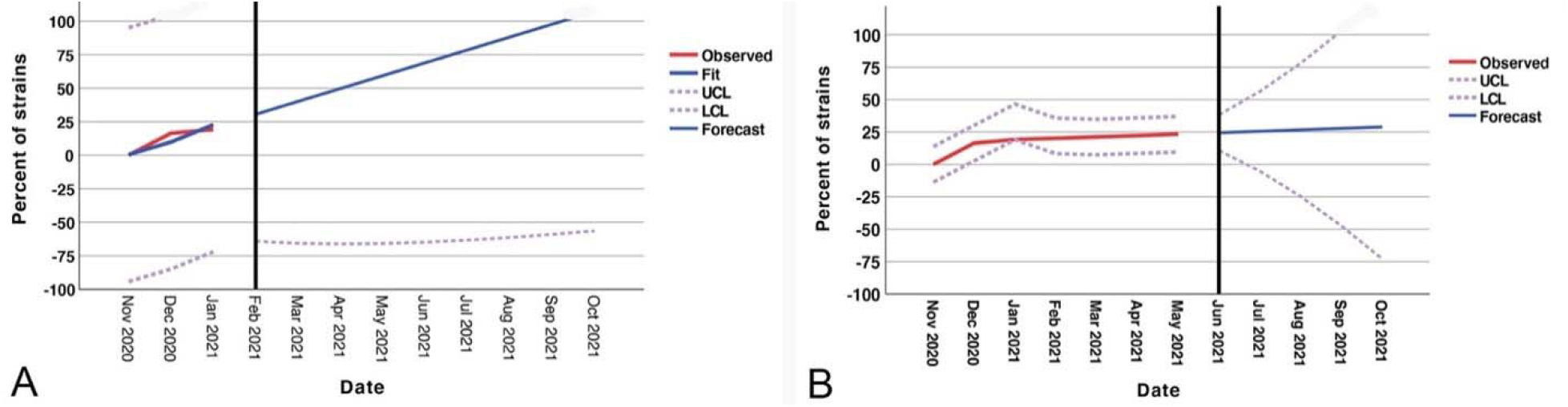
The forecast models of the proportion of SARS-CoV-2 B.1.1.7 lineage in the North American A: model 1(vaccination has no effect), B: model 2 (vaccination reduces the rate of this lineage by 70%)

Docking structure in a model interacting between SARS-CoV-2 S-RBD protein and Fab of CV30 antibody shows a rough binding affinity for the wild type S. This analysis also demonstrates overlapping between wild and mutant S-RBD proteins with human hACE-2 (Fig.3). However, the same analysis for the CV30 antibody and S-RBD of the B.1.1.7 and B.1.351 shows 8%-30% (consequently) lower binding scores in comparison to wild type as shown in Table 1. Two residues, N501Y and 484K, from the B.1.351 mutant S-RBD interact with residues from the light chain of the CV30 antibody. The aligned structure of wild type S-RBD showed that the ranking binding mode, one that corresponds to the best-scored, did not induce any significant conformational changes in the S-RBD from the CV30 antibody-bound complex. The aligned RBD had an RMSD of 2.32 Å base on docking structure. But the S-RBD mutant showed significant conformational changes by RMSD of 324.79 Å (Table 1). Three residues of B.1.1.7 (484E, 417K and 501Y) have shown hydrogen bond with hACE-2 but lower in wild S-RBD with two residues (484E and 417K) (Fig.4). Residue 417K from the wild S-RBD interacts with 52Y CV30 antibody in the domain of both chains (light and heavy). But residues 501Y and 484K mutant from B.1.351 mutant interact with light chain S52 and D70 respectively (Fig.5). We also presented docking structures of the interaction of N501Y, E484K and K417T mutations (P.1 variant) and only E484K mutation (Fig.6) in complex with a potent neutralizing CV30 antibody/hACE-2. Superposition of wild/mutant SARS-CoV-2 S-RBD protein structures also shown in Supplementary Fig 2. Summary of the findings of this study has been shown in Fig.7.

**Table 1.** Docking score of binding of S-RBD with CV30 antibody/hACE-2 from wild mutants (U.K. and South Africa).

|  | CV30 |  |  |  |  | hACE-2/Spike |  |  |  |
| --- | --- | --- | --- | --- | --- | --- | --- | --- | --- |
| Rank | *WT | B.1.1.7 | B.1.135 | P.1 | E484K | *WT | B.1.1.7 | B.1.135 | P.1 |
| Docking Score | -371.61 | -337.93 | -260.34 | -270.72 | -263.21 | -310.19 | -353.32 | -266.76 | -334.69 |
| Ligand rmsd (Å) | 2.32 | 142.44 | 324.79 | 385.41 | 74.78 | 3.27 | 0.79 | 93.91 | 0.68 |
| Score Affinity % | 100 | 90.93 | 70.05 | 72.85 | 70.82 | 100 | 113.90 | 85.99 | 107.98 |

\* Wild species data was considered with a 100% index, and data mutations were measured according to this criterion. The green bar line depicts S-RBD protein interaction with neutralizing CV30 antibody, and the red bar line depicts S-RBD protein interaction with hACE-2. WT: wild type virus.

**Figure 3.**
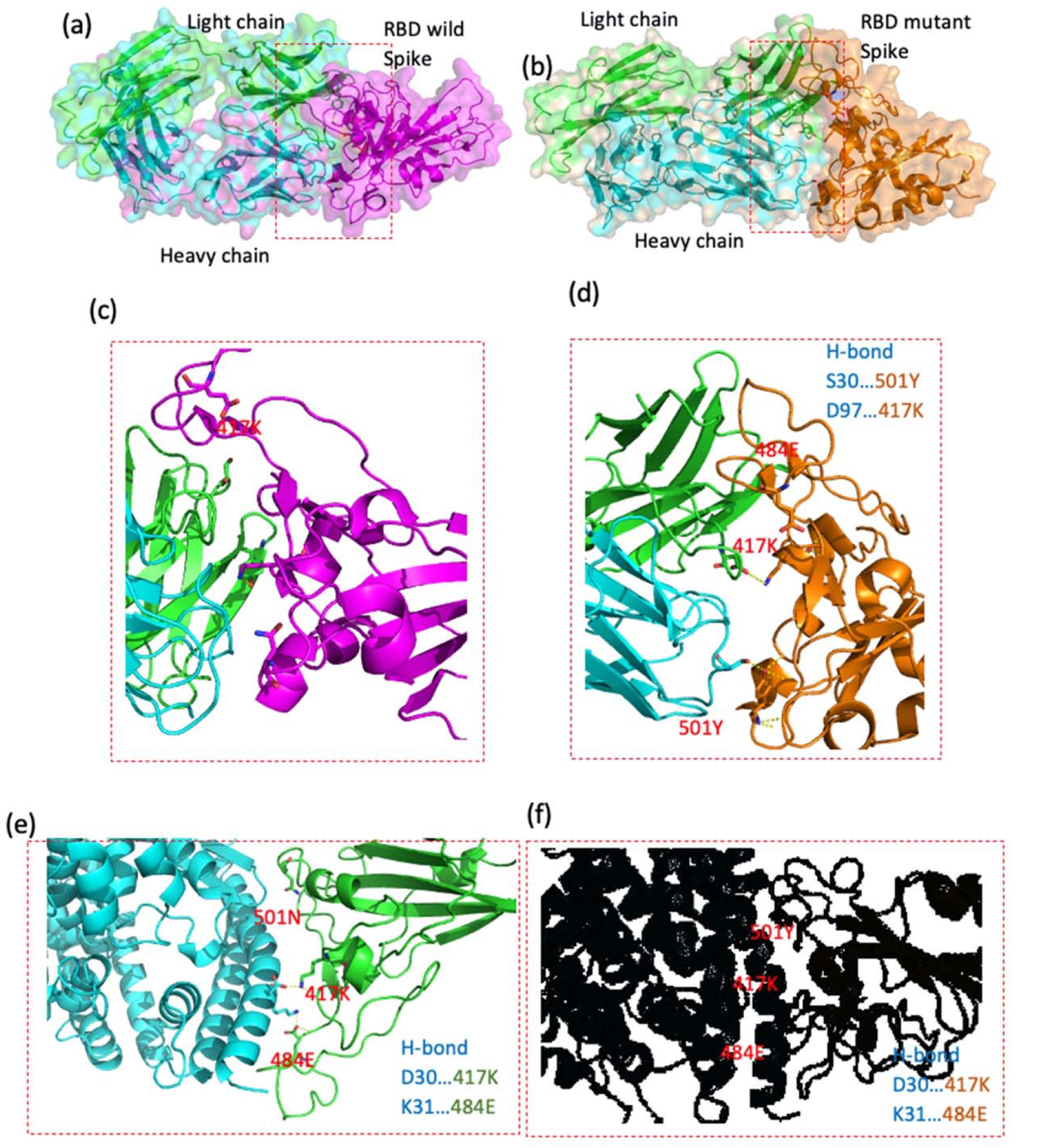
Docking structures of the interaction of SARS-CoV-2 S-RBD protein in complex with a potent neutralizing CV30 antibody. (a,c) The structure of interactions of wild S-RBD protein with neutralizing CV30 antibody. (b,d) Docking structure of interactions of mutant U.K. S-RBD protein with neutralizing CV30 antibody. (e) The structure of interactions of wild S-RBD protein with human hACE-2. (f) The structure of interactions of mutant U.K. S-RBD (N501Y) protein with human hACE-2. (The dashed line indicates the hydrogen bond).

**Figure 4.**
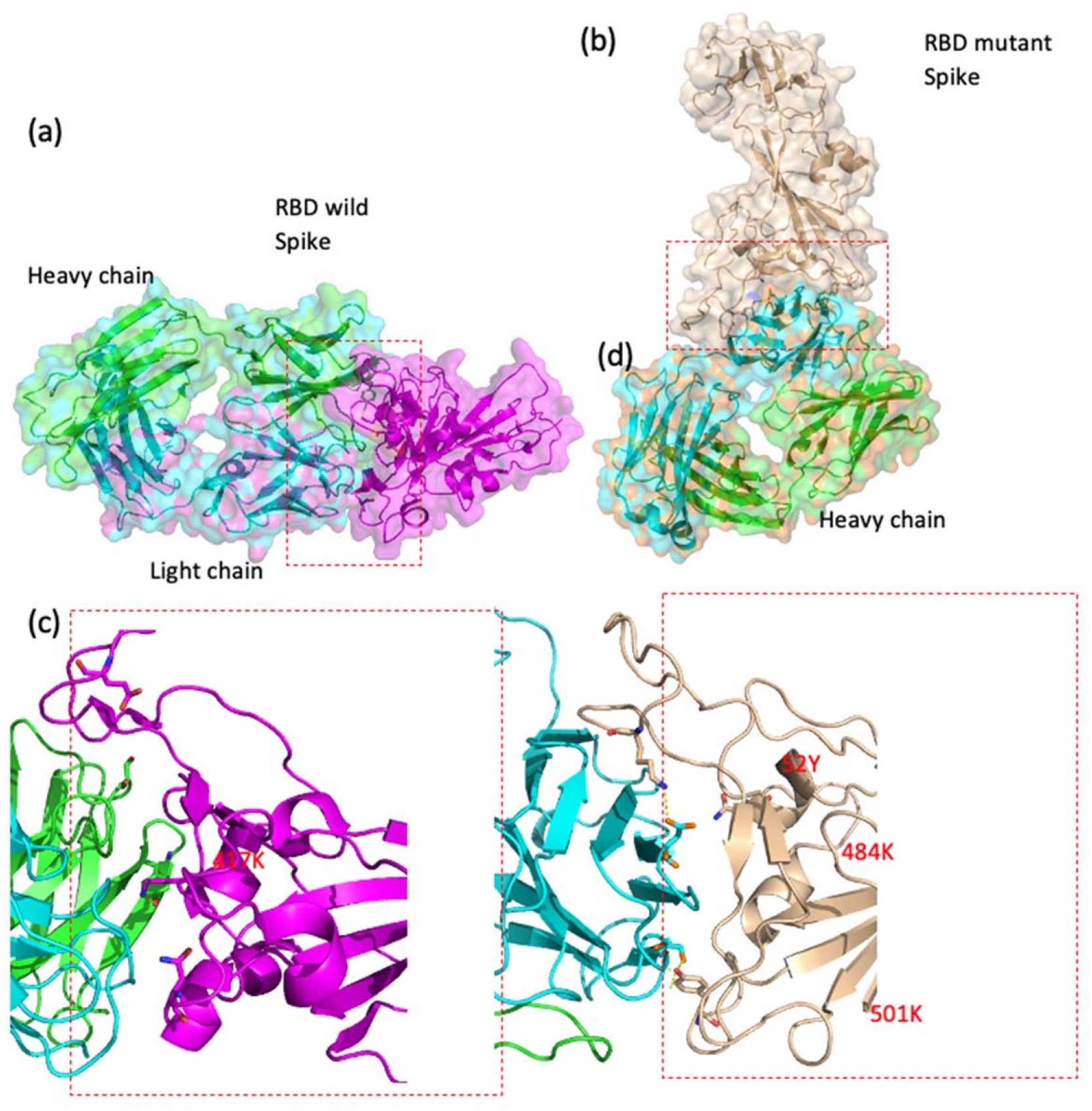
Docking structures of the interaction of SARS-CoV-2 S-RBD protein in complex with a potent neutralizing CV30 antibody. (a,c) The structure of interactions of wild S-RBD protein with neutralizing CV30 antibody. (b,d) Docking structure of interactions of mutant S-RBD protein with neutralizing CV30 antibody.

**Figure 5.**
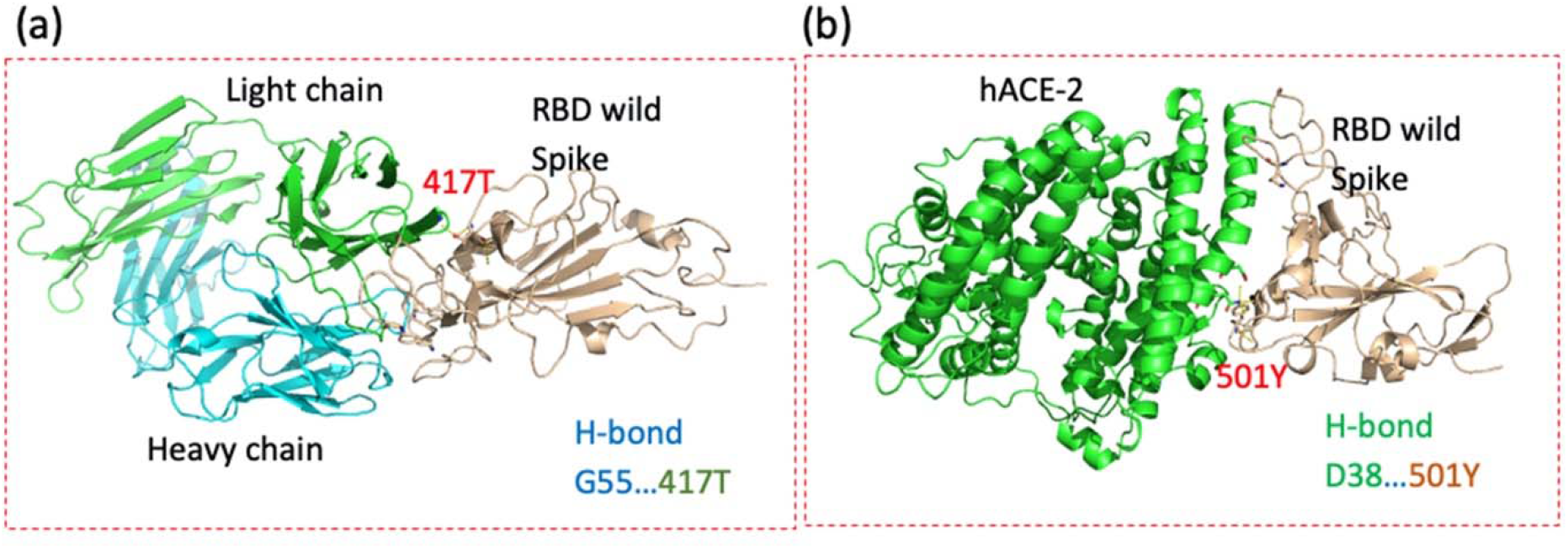
Docking structures of the interaction of P.1 mutant (N501Y, E484K and K417T) SARS-CoV-2 S-RBD protein in complex with a potent neutralizing CV30 antibody/hACE-2. (a) The structure of interactions of P.1 mutant. S-RBD protein with neutralizing Ab (b) The structure of interactions of P.1 mutant. S-RBD mutant protein with human hACE-2. (The dashed line indicates the hydrogen bond).

**Figure 6.**
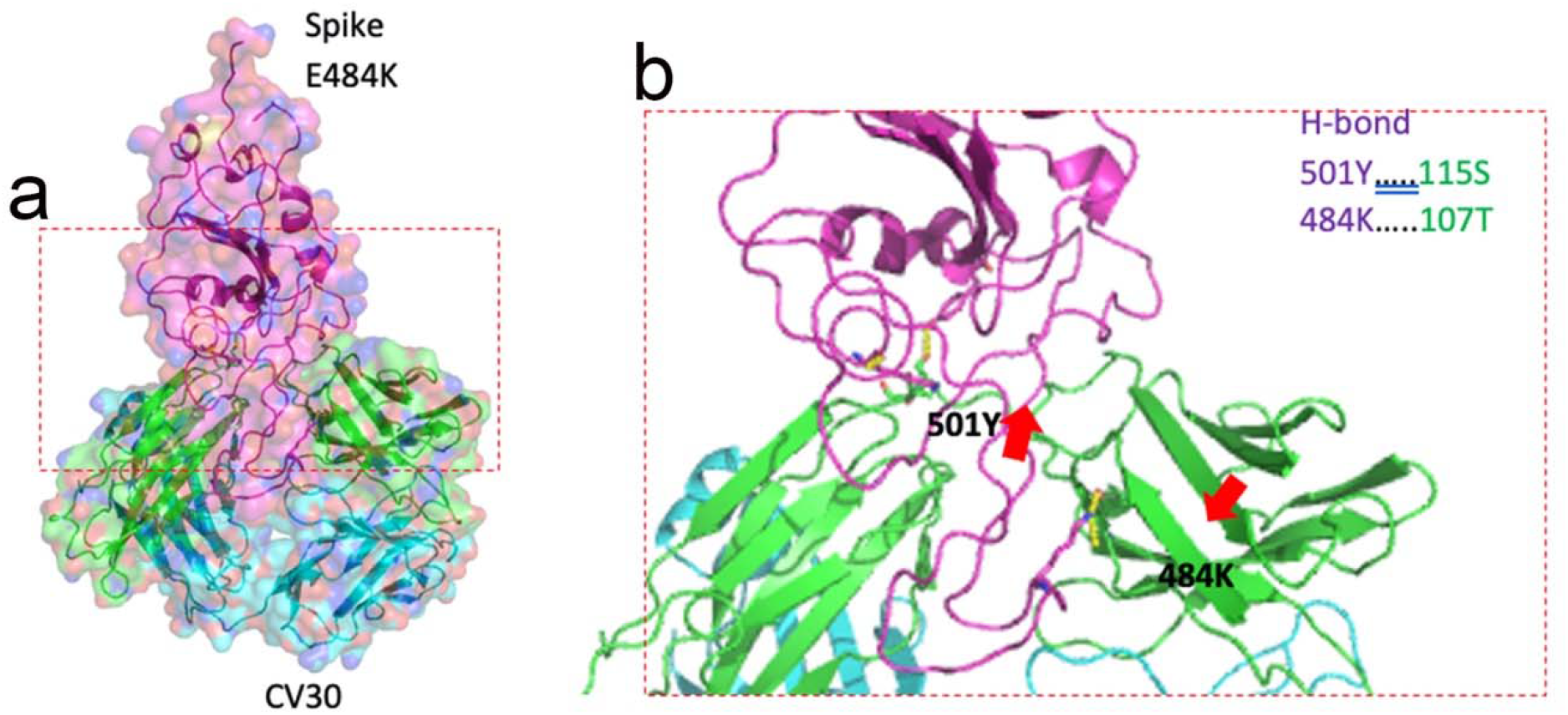
Docking structures of the interaction of E484K mutant SARS-CoV-2 S-RBD protein in complex with a potent neutralizing CV30 antibody/hACE-2 (a) surface display (b) cartoon display of maginified protein’s secondary structure. The dashed line indicates the hydrogen bond.

**Figure 7.**
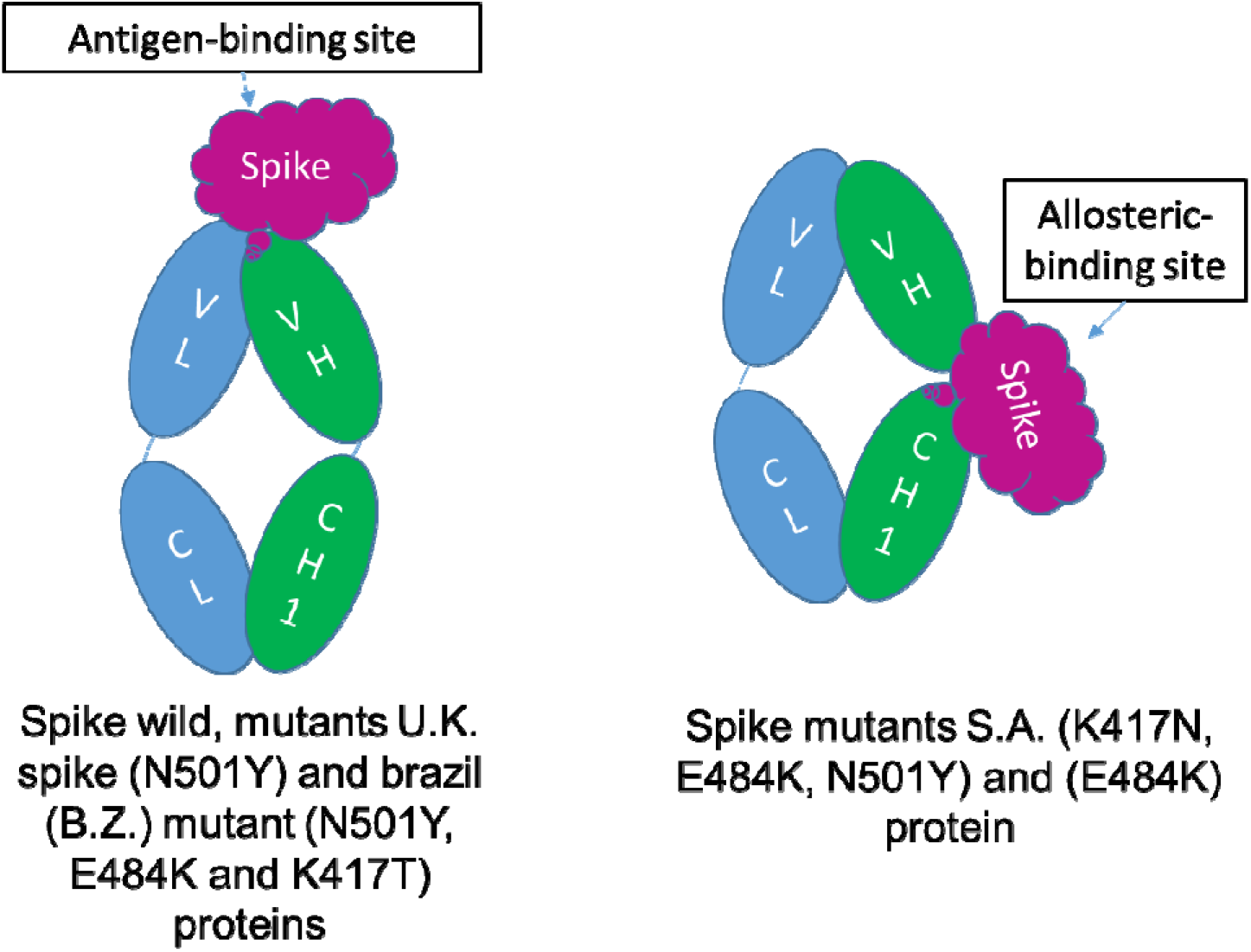
A summary of the findings of this study. The location of antiboy attachment is different btween strains.

## Discussion

The current study shows that the first mutations at positions 501 and 484 had been tracked at the same time (July 2020) from the same place (Côte d’Ivoire; West Africa). This finding indicates that the two positions (501 and 484) in S-RBD of SARS-CoV2 may have a critical role in virus adaptation and evolution during the pandemic. E484K has been reported from the South, Northeast, and North Brazilian regions in late August, 2020 by emerging of P.1 variant ^9^. Development of E484K mutation (P.1 lineage) in Brazil (Rio de Janeiro) ^9^ and B.1.351 variant in South Africa at late August ^5^, indicating that 484 position may be the top hot spots involve in infectivity, immune escape, and reinfection. N501T substitution, which probably facilitates N501Y substitution at position 501, emerged in early July 2020 at Côte d’Ivoire (west Africa) and was then detected in the U.S. in late August 2020, indicating that SARS-CoV-2 variants carrying mutations at position 501 might have circulated unnoticed even before the rapidly emerging lineages such as B.1.1.7, B.1.351 and P.1 (carrying the N501Y mutation) ^5,21^.

As of January 13, 2021, approximately 76 cases of B.1.1.7 have been detected in different states of the U.S.^18^. B.1.135 and P.1 have also been detected at the end of January 2021 ^23,24^. Our timeline analysis also confirmed that variants carrying N501Y mutation (B.1.1.7, B.1.135 and P.1) have emerged between the end of 2020 and the beginning of 2021 in the U.S (Fig.1). Analysis of SARS-CoV-2 genotypes from California revealed that a new variant called B.1.429 (CAL.20C) was first observed in July 2020 in 1 out of 1247 samples from Los Angeles County; thereafter, this variant’s prevalence has increased to 35% and 44% in California state and Southern California respectively in January 2021 ^25^. A recent study reported the presence of other important SARS-CoV-2 variants but at low rates (B.1.1.7; 23 cases, B.1.351; 2 cases, P.1; 4 cases and two closely related Cal.20C variants; B.1.429; 143 cases and B.1.427; 19 cases) among 20,453 virus specimens from COVID-19 patients dating from December 2020 through mid-February 2021 in the Houston metropolitan region ^26^. Our forecast analysis predicted that the N501Y mutant (B.1.1.7 lineage) will potentially be the most dominant strain circulating in North America in the coming months. The confirmed cases infected with the B.1.1.7 variant are rapidly increasing; 4686 confirmed B.1.1.7 variant in the U.S, alone, but the rate of other variants, such as B.1.135 (142 cases) and P.1 (27 cases) is not increasing, as rapidly (https://www.cdc.gov/coronavirus/2019-ncov/transmission/variant-cases.html, 03/11/2021).

Nelson *et al*., simulated the molecular dynamics of different mutations in the B.1.351 variant and found that the E484K mutation enhances S-RBD affinity for hACE-2 and a combination of all three mutations, E484K, K417N and N501Y, induces conformational change greater than N501Y mutant alone ^27^. However, our analysis revealed that while B.1.351 variant has ∼14% lower affinity (85.99%) to hACE-2 receptor relative to the wild type, the B.1.1.7 variant has a 14% higher affinity (113.93%) to this receptor relative to the wild type (Table 1). These discrepant results most likely occurred due to different affinity of modeled neutralizing antibodies. Notably, allosteric effects of mutant’s S-RBD variant B.1.351 with neutralizing CV30 antibody could play important role in immune evasion. Between the S-RBD protein of SARS-CoV-2 and hACE2, wild type showing that the binding free with a small RMSD but also be an important RMSD contribution to the entropy upon binding. That means that mutants S-RBD with high RMSD must overcome much more entropy penalty than wild S-RBD when binding to hACE2; energy is required for a system to change RMSD.

Both B.1.351 and B.1.1.7 variants showed a significant reduction (∼30%) of their affinity to the neutralizing antibodies relative to wild type, indicating that N501Y confers escaping ability to the new variants from the neutralizing antibodies. The B.1.351 variant could escape from neutralizing antibodies isolated from COVID19 donor plasma as well as neutralizing CV30 antibody ^28-30^. Our analysis showed the chance of escape form neutralizing antibodies could be high (∼30%) for N501Y variants. Similar to Nelson *et al*. ^27^, our analysis also showed that the combination of E484K, K417N, and N501Y results in the highest degree of conformational alterations of S-RBD when bound to neutralized CV30 antibody, compared to wild type. The variant carrying E484K (B.1.351 variant) can escape neutralization by existing first-wave anti-SARS-CoV-2 antibodies and re-infect COVID19 convalescent individuals ^27^.

Wang *et al*., showed that the mutants carrying the N501Y mutation (such as B.1.1.7) are relatively resistant to a few mAbs targeting S-RBD and not more resistant to convalescent plasma or vaccinee sera, potentially due to lack of the E484K mutation, while variants carrying mutations (such as B.1.351 which carries the E484K mutation) are resistant to multiple mAbs targeting S-RBD and are markedly more resistant to neutralization by convalescent plasma (9.4 fold) and vaccine sera (10.3-12.4 fold) ^31^. Another recent study showed that serum samples obtained from vaccinated individuals (15 participants vaccinated by Pfizer-BioNTech vaccine) efficiently neutralized wild type virus. This study also shown that neutralization of B.1.1.7 and P.1 viruses was roughly equivalent relative to neutralization of wild type but neutralization of B.1.351 virus was lower but still robust ^32^.

These data are in line with our analysis showing that emergent variants harboring mutations in S-RBD (E484K, K417N, and N501Y) will pose significant challenges for mAb therapy and threaten the protective efficacy of current vaccines. Our data suggest that a 30% reduction in antibody affinity to mutant strains would be translatable to up to a 9-fold increase in viral replication rate. Taken together, these data highlight the prospect of reinfection with N501Y mutants as antigenically distinct variants and may foreshadow reduced efficacy of current S-RBD -based vaccines in the near future. The combination of E484K, K417N, and N501Y results in the highest degree of conformational alterations in the interaction between S-RBD and human hACE-2, compared to either E484K or N501Y alone ^27^. Currently, three vaccines are authorized and recommended to prevent COVID-19; Pfizer-BioNTech, Moderna, and Janssen & Janssen (J&J). All three vaccines are designed to encode S-RBD ^33-35^. Moderna’s COVID-19 vaccine produced neutralizing titers against all key emerging variants tested, including B.1.1.7 and B.1.351, with no significant impact on neutralizing titers against the B.1.1.7 variant but a six-fold reduction in neutralizing titers against the B.1.351 variant relative to prior variants ^36^. Despite this reduction, neutralizing titer levels with B.1.351 remain above levels that are expected to be protective ^36^. Another study also shown that the neutralizing activity against B.1.1.7 was essentially unchanged, but was significantly lower against B.1.351 (12.4 fold, Moderna; 10.3 fold, Pfizer) ^31^, but no data available for J&J as this vaccine was approved later. The Oxford Covid-19 vaccine, which has been approved in the U.K., has also shown a substantially reduced neutralization effect against B.1.351 variant compared to wild type ^37^, but more studies are already underway at the University of Oxford for further evaluation of new mutations on Oxford Covid-19 vaccine efficacy.

## Conclusions

As the proportion of B.1.1.7 strain which has more affinity for hACE-2 receptor has been increasing in the U.S., our concerns about loss of vaccination efficiency have been growing. Our computation suggests that the docking interaction score between the S-RBD and hACE2 proteins were less favorable for the B.1.351 (K417N, E484K, and N501Y) than for the B.1.1.7 and P.1 strains as compared to the reference S-RBD. Our results also indicated a weak interaction between mutants S-RBD of B.1.1.7, B.1.351 and P.1 with the neutralizing CV30 antibody and could lead to poor immunity. Thus, it is expected that B.1.351 will decrease affinity to the CV30 antibody by ability allosteric binding. RMSD of the S-RBD complex by ACE2 from the B.1.1.7 and P.1 variants and wild type are very low, suggesting that they should have a similar topology. The present study is entirely in silico work and should be validated by future studies before conducting trials in humans.

## Method

### Time series forecasting

SARS-CoV-2 genomic sequence data were gathered from the Nextstrain global database (https://nextstrain.org/), and mutation site numbering and genome structure was done using the Wuhan-Hu-1/2019 genomic sequence as reference (wild type). The database created by the Global Initiative on Sharing All Influenza Data (GISAID) and the analysis tool Nextstrain have enabled the analysis of thousands of whole-genome sequences of SARS-CoV-2 ^38^. Our analysis was performed only for mutations involving the S-RBD domain (see supplementary Figure 1 for more details). The Nextstrain data bank groups the isolates by regions. Once prevalent mutations were identified, the percentages of mutant genomes were calculated from the total regional subsample of isolates for various time periods between December 2019 to the end of January 2021. In addition, separate percentages were calculated from the date of the first appearance of each mutation to the present. Data from North America has been updated through January 9, 2021.

A time series forecast was developed for the SARS-CoV-2 B.1.1.7 variant in North America. The current U.S. percentage of B.1.1.7 was calculated using Nextstrain. The first model was developed with an assumption that vaccination at the current rate will not affect the proportion of the B.1.1.7 mutant variant in the North America and assuming equal and sustained rates of infectivity (R 1.1). We then conducted another forecasting model with the assumption of 100% vaccination coverage in North America. Although the effectiveness of the COVID19 vaccine and time duration of protection remains to be determined, we assumed 100% efficacy, with a duration of 12 months of protection.

### Ethical Approval statement

We confirm that all experimental protocols were approved by Nextstrain. Authors confirm that all methods were carried out in accordance with relevant guidelines and regulations.

### Construction of the computational model

Atom coordinates of wild S-RBD in complex with neutralizing CV30 antibody were extracted from the crystallography structure of the Protein Data Bank (RCSB PDB) with the access code 6XE1 ^39^. The neutralizing monoclonal CV30 antibody was isolated from a patient infected with SARS-CoV-2, in complex with the receptor S-RBD ^40^. We used CV30 antibody in our modeling as its binding to the concave ACE2 binding epitope of the S-RBD has previously been well characterized ^39^. We obtained hACE2 from electron microscopy of 6CS2 for SARS S Glycoprotein-hACE2 complex ^23^. The crystal structure of the N501Y mutant S-RBD of SARS-CoV-2 Spike was obtained from RCSB with the access code 7NEG. The tertiary structure of the S-RBD mutant South Africa and Brazil mutants were predicted using the Phyre2 server ^24^. HDOCK Server ^41^ (http://hdock.phys.hust.edu.cn/) was utilized to examine protein-protein interaction based on a hybrid algorithm of template-based modeling into free docking with default parameters. To prepare structures as input files for a docking run, all water molecules and solvent molecules were removed from the proteins. All of the structures were visualized using PYMOL Chimera software version 1 ^6^. The calculation procedure was almost the same as that in our previous work ^42-44^ with neutralizing, antibodies were separated from SARS-CoV-2 S-RBD into individual files for docking simulation. The root-mean-square deviation (RMSD) was calculated to evaluate the change in the geometric structure of S-RBD after docking by using backbone atoms. RMSD was used to quantify the similarity between two superimposed atomic coordinates of the reference structure (PDB structure) and the post-structure complex with receptor molecules. The RMSD was calculated to evaluate changes in the geometric structure of the spike protein using spinal atoms.

## Data Availability

All data generated and analyzed during this study are included in this article [and its supplementary information files].

## Abbreviations

SARS-CoV1: Severe acute respiratory syndrome coronavirus 1
MERS-CoV: Middle East respiratory syndrome coronavirus
S-RBD: Spike receptor-binding domain
hACE-2: Human angiotensin-converting enzyme 2
U.S: United State
U.K: United Kingdom
COVID19: Coronavirus disease 2019
CDC: Centers for Disease Control
GISAID: Global Initiative on Sharing All Influenza Data
RCSB PDB: Protein Data Bank
RMSD: Root-mean-square deviation

## Ethics approval and consent to participate

Not applicable. Nextstrain is an all-source code is freely available under the terms of the GNU Affero General Public License. Nextstrain is supported by NIH for research on pathogens including SARS-CoV2. We have accessed to de-identifiable data.

## Consent for publication

Not applicable

## Author’s Contributions

EQ contributed to the acquisition, analysis, and interpretation of data, as well as drafting the manuscript. MV contributed to computational analysis, drafting the manuscript. AHS contributed to the draft revision. MM contributed to the conception of study, the acquisition, analysis, draft revision, and supervision.

## Acknowledgment

Authors would like to thank April Mann for critical review and edits

