## Supplementary file for "A time series forecasting of the proportion of SARS-CoV-2 N501Y lineage in North America"

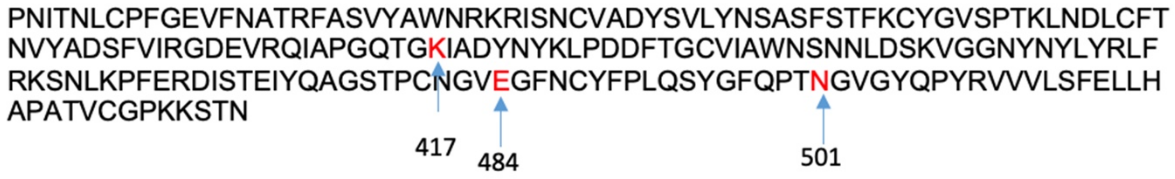


**Supplementary Figure1**. Sequence alignment of S-RBD SARS-CoV-2 genes with mutations K417N, E484K, N501Y

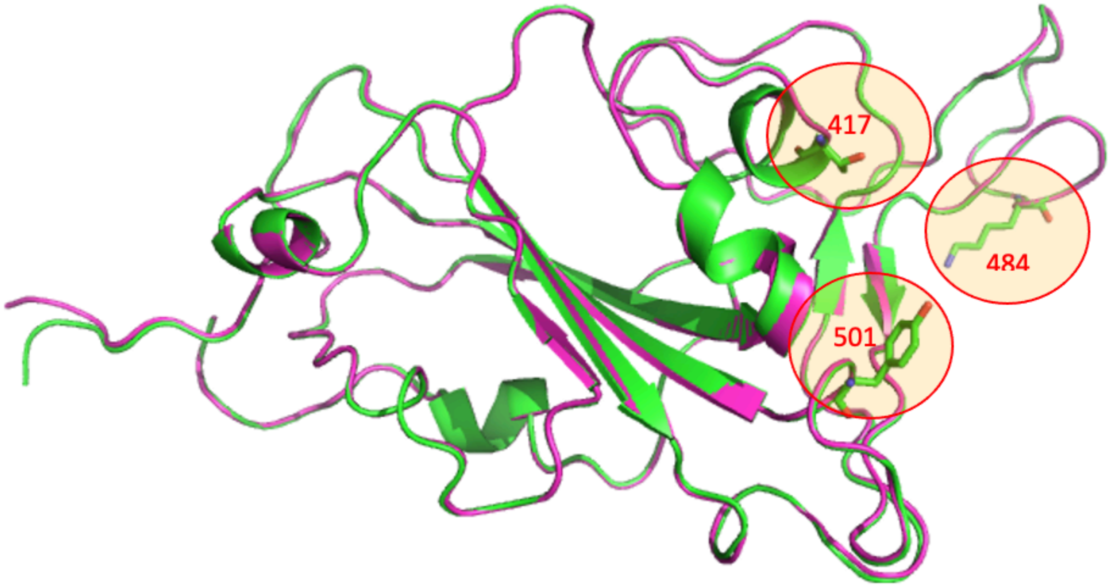


**Supplementary Figure 2.** Superposition of wild/mutant SARS-CoV-2 S-RBD protein structures. The red rings in the photo indicate the three mutation points in the S-RBD that are in the flexible coil region. The structure displayed in purple indicates wild and the green color indicates mutated.
